# ENVIRONMENTAL SAFETY EVALUATION OF THE PROTECTION AND ISOLATION SYSTEM FOR PATIENTS WITH COVID-19

**DOI:** 10.1101/2020.06.04.20122838

**Authors:** Claudio Almeida Quadros, Maria Carolina Bezerra Di Medeiros Leal, Almeida Baptista Sobrinho Carlos de, Carolina Kymie Vasques Nonaka, Bruno Solano de Freitas Souza, Juliana Cristina Milan-Mattos, Aparecida Maria Catai, Valéria Amorim Pires Di Lorenzo, Antonio Gilberto Ferreira

## Abstract

**Background:** SARS-CoV-2 has high transmissibility through respiratory droplets and aerosol, making COVID-19 a worldwide pandemic. In its severe form, patients progress to respiratory failure. Non-invasive mechanical ventilation restrictions and early orotracheal intubation have collapsed health systems due to insufficient intensive care unit beds and mechanical ventilators. COVID-19 dedicated healthcare professionals have high infection rates. This publication describes experimental testing of the Protection and Isolation System for Patients with COVID-19 (PISP/COVID-19).

**Method:** PISP/COVID-19 is a disposable transparent polyethylene plastic that covers the patient’s entire hospital bed, with its internal air aspirated by the hospital’s vacuum system attached to a microparticle filter. Experiments validated PISP/COVID-19’s ability to block aerosolized microparticles dissemination. Caffeine was used as a molecular marker, with leakage evaluation through sensors analysis using nuclear magnetic resonance spectroscopy. The biological marker was synthetic SARS-CoV-2 RNA, using Reverse Transcription Polymerase Chain Reaction (RT-PCR) as the detection method.

**Results:** PISP/COVID-19 was effective against molecular and biological markers environmental dispersion in simulations of non-invasive ventilation, high-flow nasal cannula oxygen and mechanical ventilation isolation. Caffeine was not detected in any of the sensors positioned at points outside the PISP/COVID-19. The ability of PISP/COVID-19 to retain virus particles and protect the surrounding environment was confirmed by detection and gradients quantification of synthetic SARS-CoV-2 RNA by RT-PCR.

**Conclusion:** PISP/COVID-19 was effective in the retention of the molecular and biological markers in all tested simulations. Considering the current pandemic, PISP/COVID-19 might increase the use of non-invasive ventilation, high-flow nasal cannula oxygen and provide additional protection to healthcare professionals.

## 1. Background

COVID-19 is a disease caused by the new coronavirus (SARS-CoV-2) with varied clinical manifestations, ranging from flu-like symptoms to severe pneumonia.^1–4^ SARS-CoV-2 has high transmissibility, mainly from person-to-person, through respiratory droplets produced by expiration, coughing or sneezing from people who are infected,^1,3,5^ which has led to its rapid global dissemination.^4,6^ In March 2020, three months after the first case of COVID-19, the World Health Organization (WHO) declared it as a pandemic.^2,7^

Hospital contamination by dispersion of respiratory droplets with SARS-CoV-2 made health professionals’ occupational safety a challenge.^6–8^ Studies indicate that viral particles in the environment remain viable for hours,^4,8^ which explains the high incidence of contamination of health teams. Similar to the transmission mode of the current pandemic, half of all SARS-CoV-1 cases observed were caused by nosocomial contamination to health professionals during the outbreak in Canada in 2003.^5^

Treatment guidelines for patients with COVID-19 who progress to respiratory failure indicate early orotracheal intubation and maintenance on mechanical ventilation,^4,6^ in addition to isolation.^4,6,9,10^ The guidelines for early intubation has caused high demand for hospitalization in the Intensive Care Unit (ICU) and the need for mechanical ventilators therefore collapsing health systems.^11^

Strict requirements for non-invasive ventilation in COVID-19 patients has made it become an unfeasible medical treatment.^5,9,12^ Non-invasive ventilation (NIV) even when indicated, is not performed in order to avoid contamination within the hospitals from viral particles in aerosols, which are generated by the positive pressure of ventilation in face masks and high oxygen flow.^5,9,12^ Patients affected by acute respiratory syndromes during the 2003 (SARS) and 2009 (H1N1 virus) epidemics that were submitted to NIV had shorter hospital stays and lower mortality rates.^13^ They presented similar results as hospitalized patients who did not need ventilatory support.^13^ The benefit of NIV recommendation can also be seen in data that reveals that the mortality rate of patients for whom NIV support did not work and further needed mechanical ventilation was similar to the ones who needed mechanical therapy as the initial treatment of respiratory acute failure.^13^

Given this scenario, this article proposes a simple and easy implementation system for the isolation of patients with COVID-19, avoiding contamination within the hospital environment and thus increasing the protection of health professionals and other hospitalized patients. The system is called: Protection and Isolation System of Patients with COVID-19 (PISP/COVID-19). The present study aims at evaluating the effectiveness of the proposed system in retaining the aerosolized molecular marker and synthetic SARSCoV-2 RNA, in simulations with NIV, high-flow nasal cannula oxygen and isolation of patients under mechanical ventilation. Simulations included extreme situations of aerosol release. The evaluations were performed through environmental chemical and molecular monitoring by analytical analysis using nuclear magnetic resonance spectroscopy (NMR) and Real Time Reverse Transcription Polymerase Chain Reaction (RT-PCR).^1^

## 2. Materials and Methods

### 2.1 Description of the Protection and Isolation System of Patients with COVID-19 (PISP/COVID-19)

PISP/COVID-19 has two security features: a physical barrier between the patient and the external environment, and an air suction system with a micro-particle filter. The physical barrier is a disposable transparent polyethylene plastic cover (5 × 4 × 0.001m) from Bhiosupply®. In order to keep the plastic cover elevated, a sterilizable, stainless steel and polycarbonate structure was designed, allowing the system to be dynamic and adaptable to any type of hospital bed (Bhiosupply®). The plastic cover is attached to the hospital bed by an elastic band made of polyester and elastane (figure 1). The micro-particle filter used in the air suction inside PISP/COVID-19 has 0.1–0.2 µ porous membrane (from Bhiocap®) and is connected to the hospital vacuum system or it can also be connected to a pump machine with suction pressure of 40 l/min.

**Figure 1.**
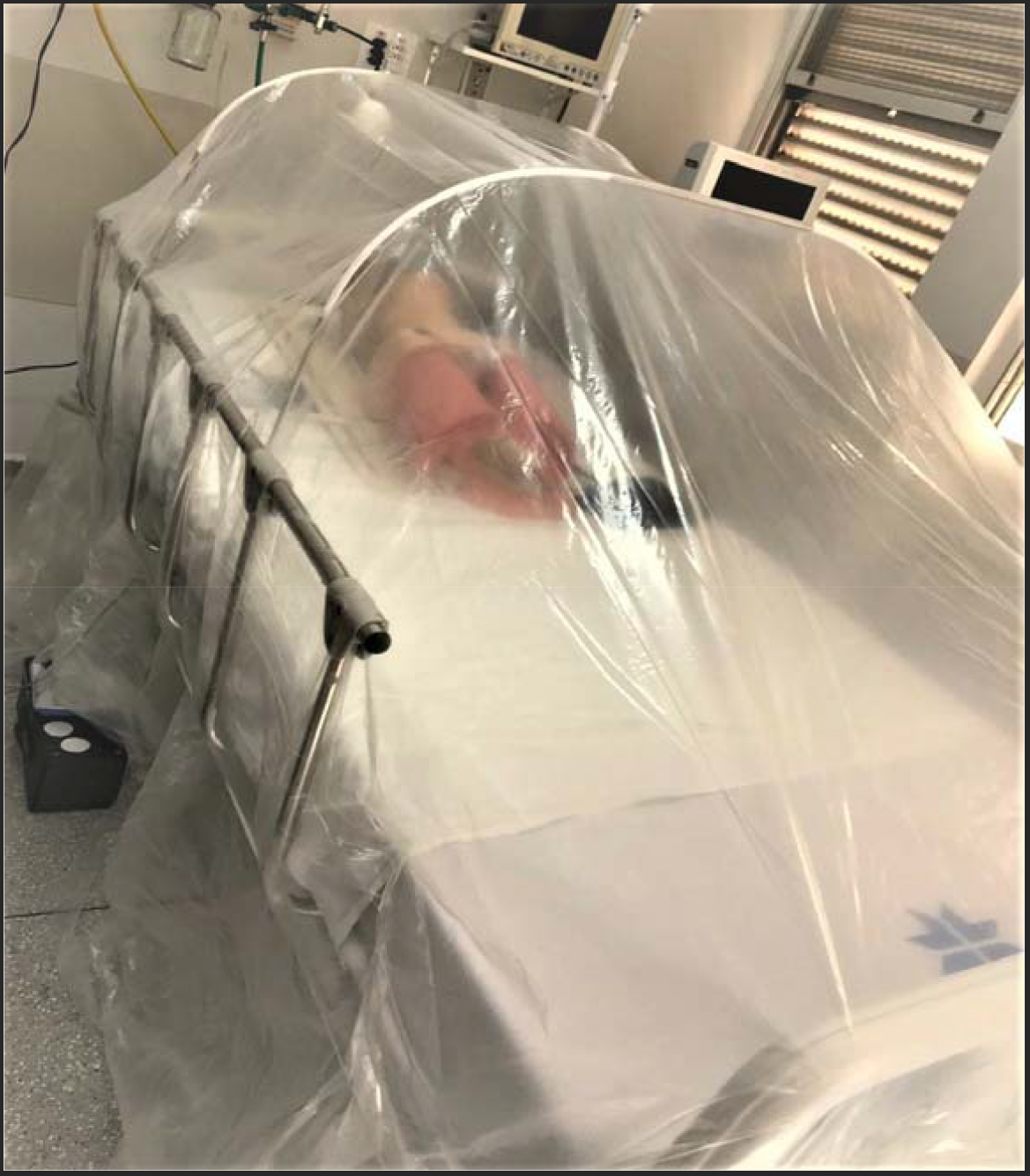
PISP/COVID-19.

### 2.2 Chemical monitoring of the molecular marker

Chemical monitoring inside and outside the PISP/COVID-19 was performed using strategically positioned pairs of nitrocellulose discs (NC 47 mm 8.0µ) that were used as sensors to detect the molecular marker (figure 2). Caffeine was used as a molecular marker (1,3,7-trimethylpurine-2,6-dione) being sprayed (1% w/v solution) inside PISP/COVID-19 through two different methods: first, using a Bio-Inject^®^ pump with an average pressure of 200 psi, flux of 2 ml/s, volume of 10 ml, and a second simulation mode, using a nebulizer machine. At the end of each experiment, the sensor discs were removed and put inside a 5 ml Eppendorf tube protected from the light for further analyses. The PISP/COVID-19 was discharged. The room and all non-disposable items were cleaned.

**Figure 2.**
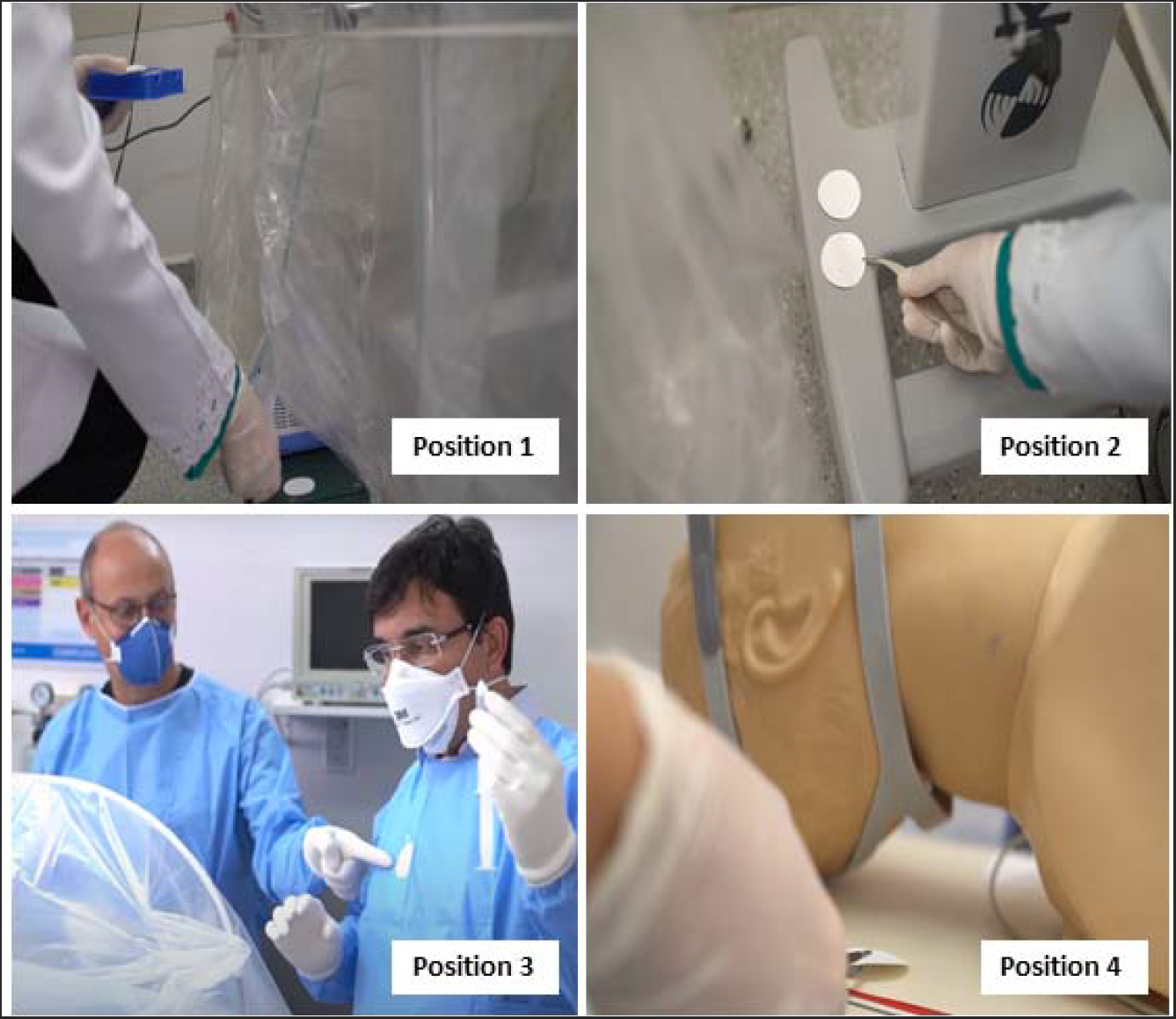
Position of nitrocellulose sensor discs: 1) the entryway of the UCI monitoring wires and respiratory tubes; 2) the entrance to the intravenous therapy tube; 3) on the health professional’s chest; 4) inside the PIS/COVID-19.

### 2.3 ^1^H NMR analysis of the nitrocellulose discs

In the Eppendorf flask with the sensor discs, 0.6 ml of D_2_O solution containing 0.2% TSP-d4 [3-(trimethylsilyl) propionate-2,2,3,3 -d4 sodium] for internal reference or H_2_O (milli-Q^®^ grade) was evaluated and sonicated for 5 min. The extracted solution was then analyzed by hydrogen nuclear magnetic resonance (^1^H NMR).

All the ^1^H NMR experiments were carried out in a 14.1 Tesla (600 MHz for hydrogen frequency) Bruker equipment, model AVANCE III, using a 5 mm TXI cryo-probe, maintaining the sample temperature at 298K during the whole experiment and in the pre-saturation of the HDO signal, using a pulse sequence with a continuous wave pre-saturation and gradient field. The acquisition parameters were: acquisition time (AQ = 4.18s), relaxation delay (d1 = 1s), spectral width (SWH = 7837 Hz), number of scans (ns = 128), received gain (rg = 128) and pulse duration (p1 = 7.7 µs). The spectra were processed using a TopSpin^®^ software (Bruker version 3.5 pl7) without an apodization function. To verify the limit of detection in the ^1^H NMR technique, a water caffeine solution from 1% to 0.00001% w/v, which corresponds to minimum 1 ppm was prepared, in the same acquisition and processing condition of the spectra described above.

### 2.4 Production of synthetic SARS-CoV-2 RNA and RT-PCR experiments

Sequences of the amplification sites of SARS-CoV-2 genes E, RdRp and N along with the T7 promoter sequence were synthetized as duplex DNA oligonucleotides and in vitro transcription was performed by using the commercially available kit T7 RiboMAX™ Express Large Scale RNA Production System (Promega, Madison, WI, USA), following the manufacturer’s instructions. The synthetic RNA transcripts were mixed in 15 mL NaCL 0,9% solution at the following concentrations: 10^10^ RNA copies/ml for E gene and 10^7^ RNA copies/ml for N and RdRp genes. Detection was performed by RT-PCR using a commercially available multiplex RT-PCR kit (Allplex™ 2019-nCoV Assay – Seegene, Seoul, South Korea), following the manufacturer’s instructions. RT-PCR was performed in a 7500 Fast Real Time PCR equipment (ThermoFisher Scientific, Waltham, MA, USA) and 45 amplification cycles were performed. The cycle threshold (Ct) results obtained for each sample tested are presented.

### 2.4 Simulation procedure of NIV

A human mannequin with a respiratory dynamics simulator and airway management model (Laerdal^®^) was placed in a hospital bed and covered with PISP/COVID-19 in all the experiments. In order to simulate the real patient’s needs in case of COVID-19, the human mannequin had a tube attached to simulate intravenous therapy, wires from the electronic UCI monitors and tubes from the NIV system. Non-invasive positive pressure ventilation was delivered to the face mask, with an inspiratory positive airway pressure set at 20 cm H_2_0 and expiratory positive pressure maintained at 5 cm H_2_0, in a continuous setting of 14 breaths/minute. In the first NIV experiment, the spray of caffeine solution (1% in D_2_O) was sprinkled into the PISP/COVID-19 with an average pressure of 200 psi, flux of 2 ml/s, volume of 10 ml for a period of 90 minutes. The suction was done through a vacuum pump, with a suction flow of 40 l/min, throughout the period. In the second simulation, the caffeine solution was sprayed using a medical nebulizer machine for 90 minutes to achieve the most realistic situation as in a physiological condition, and the vacuum hospital’s pump was used. In both simulations the internal environment of the PISP/COVID-19 system was saturated with droplets formed by aerosols. After 90 minutes, the sensor discs and the micro-particle filters were removed for chemical analysis. In the fourth experiment, 10^10^ RNA copies/ml for E gene, 10^7^ RNA copies/ml of N and RdRp genes, in 15 ml NaCL 0,9% solution, were sprayed inside PISP/COVID-19 with a nebulizer machine and internal suction was maintained using the vacuum hospital’s pump. Samples inside and outside PISP/COVID-19 were collected for RT-PCR analysis, in the same positions of the sensor disks used for detection of the molecular marker, after 60 minuts.

### 2.5 Simulation of high-flow nasal cannula oxygen

The same setting was established for high-flow nasal cannula oxygen simulation. Nasal cannula was positioned in the airway management model (Laerdal^®^) and oxygen flow was set in 80 l/minute. A nebulizer machine sprayed a 15 ml NaCL 0,9% solution with 10^10^ RNA copies/ml for E gene, 10^7^ RNA copies/ml of N and RdRp genes inside PISP/COVID-19, that had its suction maintained using the vacuum hospital’s pump. After 60 minutes, samples inside and outside PISP/COVID-19 were collected for RTPCR analysis, in the same positions described above.

### 2.6 Simulation of patient’s isolation for a 6-hour period

PISP/COVID-19 was assembled with the same details as described in the previous section. Aerosolization of the 1% caffeine solution was performed using a pump delivering 200 psi of pressure, flux of 2 ml/s, volume of 10 ml, in 120 minutes intervals, with a continuous suction pump flow of 40 l/min. Another simulation was performed using continuous nebulization as described in section 2.2 and the PISP/COVID-19 suction was performed using the hospital’s vacuum pump. The nitrocellulose sensor discs were collected every two hours.

## 3. Results and Discussion

Simulations concluded that PISP/COVID-19 is effective in the restraint of the molecular marker (caffeine) without external contamination during and after the performed simulations, using the evaluation of sensor discs with the ^1^H NMR technique, in the established detection limit of the NMR, which for this experiment was the sensor discs contamination of 1 ppm of caffeine. The results obtained in the experiment were similar when high level aerosolization pressure was delivered, with an average pressure of 200 psi, or when aerosolization was performed with a nebulizer machine. No molecular marker was obtained in any of the PISP/COVID-19 external sensor discs.

Spectrum A, in figure 3, demonstrates the standard ^1^H spectra of caffeine solution with a typical sign of this substance. It is possible to observe the chemical shift (δ) of the methyl groups region (δ = 3.28; 3.45; 3.91 ppm) and the chemical shift of the purine hydrogen (δ 7.88 ppm). The same pattern is observed in the yellow line of spectrum D, regarding the discs positioned inside the PISP/COVID-19, that were the positive control sensors. However, in spectra B and C, corresponding to the right and left sides of the hospital bed, near the tubes and wires described in section 2.4, they did not display caffeine molecules. This shows that the system was efficient because it was not possible to observe caffeine in considerable fragile points outside PISP/COVID-19.

**Figure 3.**
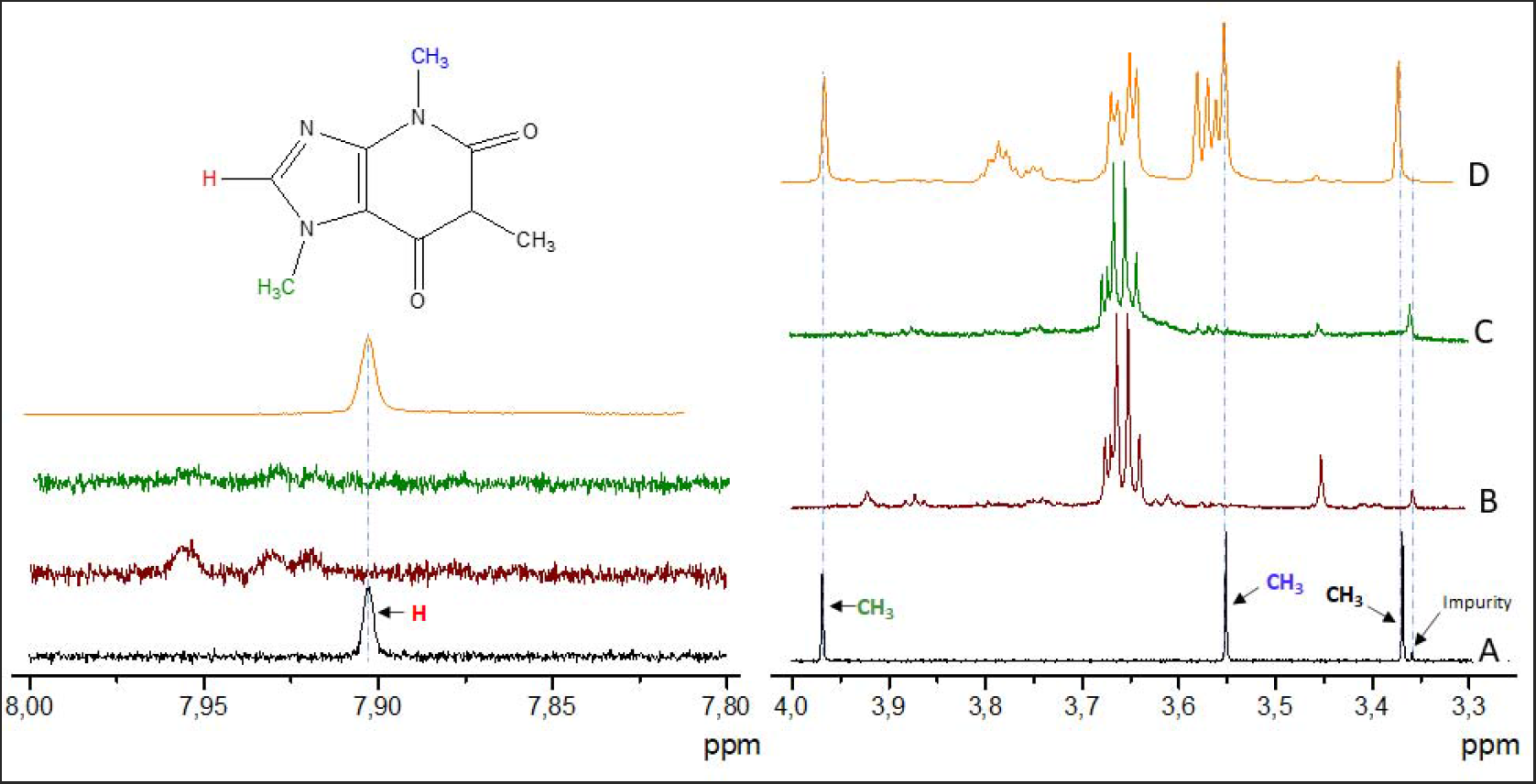
^1^H NMR spectra in D_2_O for the NIV simulation. A) caffeine solution 0.001%, B) extract from nitrocellulose sensor disc in position 1, C) extract from nitrocellulose sensor disc in position 2, D) extract from nitrocellulose sensor disc inside the PIS/COVID-19 cover.

The ^1^H NMR spectra analysis performed in the sensor discs, when the simulated patient stayed for a period of 6 hours, with the caffeine solution sprayed continually inside PIS/COVID-19, is presented in figure 4. At the end of the 6 hour period, the following sensor discs were analyzed: Positions: 1 (spectrum B) located in the entrance of the intravenous therapy tube, 2 (spectrum C) located in the entrance of the UCI monitoring wires, and 3 (spectrum D) located in the entrance of respiratory mechanic ventilator tubes. Caffeine molecules were not found on the sensor discs in any of these positions. It was only detected in position 4 (spectrum E) which corresponds to the sensor discs inside the PIS/COVID-19.

**Figure 4.**
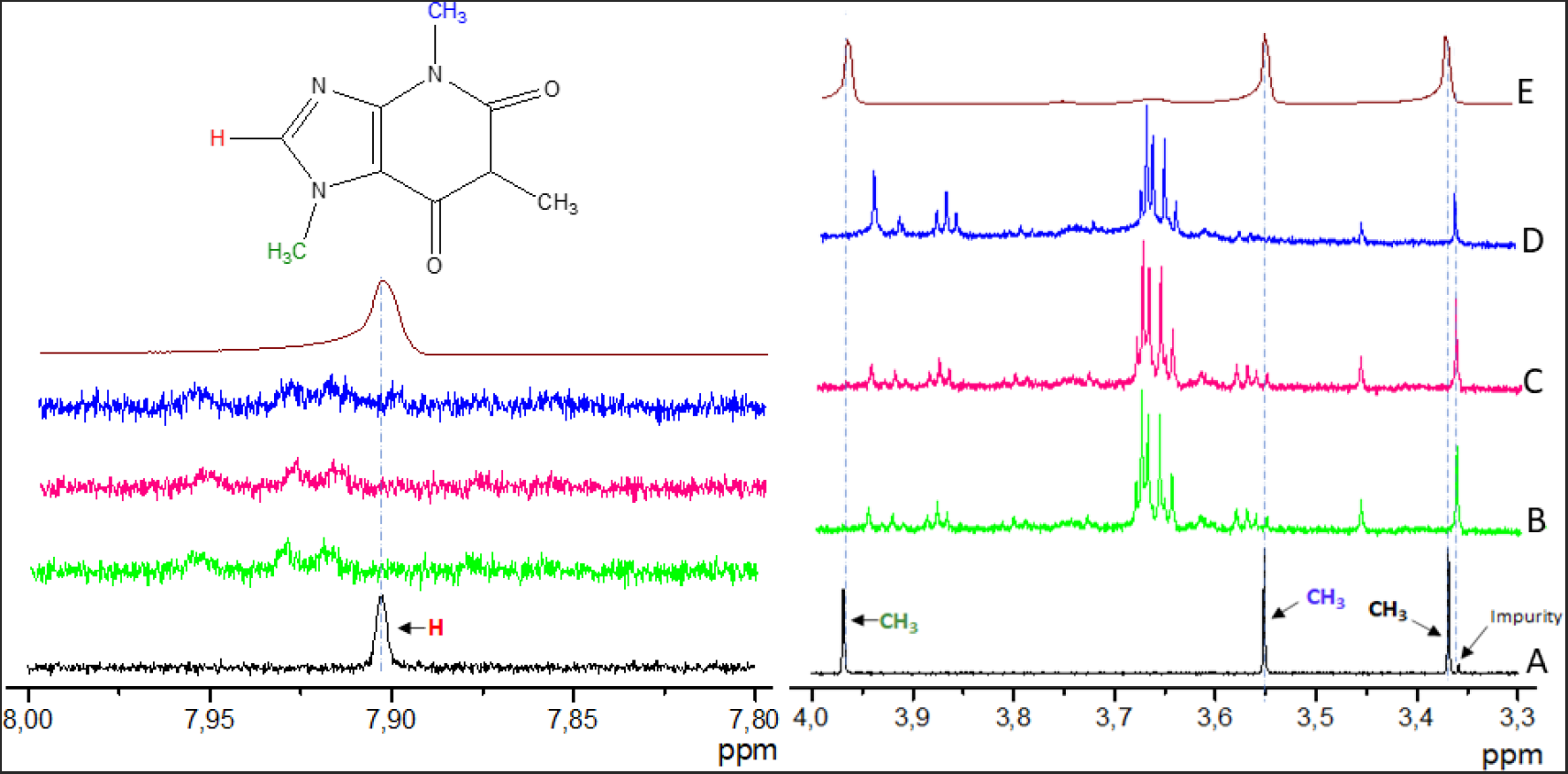
^1^H NMR spectra in D_2_O simulation during a continuous 6-hour period of patient isolation under mechanical ventilation. A) caffeine solution 0.001%, B) extract from nitrocellulose sensor disc in position 1, C) extract from nitrocellulose sensor disc in position 2, D) extract from nitrocellulose sensor disc in position 3, E) extract from the nitrocellulose sensor disc inside the PIS/COVID-19.

With the analytical findings of the isolation efficacy of the PIS/COVID-19, through NMR analysis, using caffeine as a molecular marker, the next step was naturally to evaluate its efficacy using synthetic viral particles, approaching clinical reality to COVID-19 patients. Detection was performed by Real Time PCR, in which amplification of the nucleic acid is detected by accumulation of a fluorescent signal, giving a Cycle threshold (Ct) value, which is inversely proportional to the amount of target nucleic acid in the sample

As can be seen in figure 6, the contamination of the external environment of PIS/COVID-19 was imperceptible when 10^7^ synthetic SARS-CoV-2 RNA copies/ml were used during aerosolization. Viral RNA was detected in samples taken from positions identified as number 10 (internal surface #2; plastic cover), position 11 (mannequin) and 12 (HEPA filter) for N gene and in samples taken from positions 11 and 12 for RdRp gene (figure 5A). In an extreme situation of viral particle concentration, using 10^10^ synthetic SARS-CoV-2 RNA per mL, in a 15 mL solution, viral RNA (E gene) was detected in all analyzed samples, but a scattering gradient is observed where the concentration is much higher inside the PIS/COVID-19 and gradually reduces in the external environment, as demonstrated by Ct values of RTPCR analysis (figure 5B).

**Figure 5.**
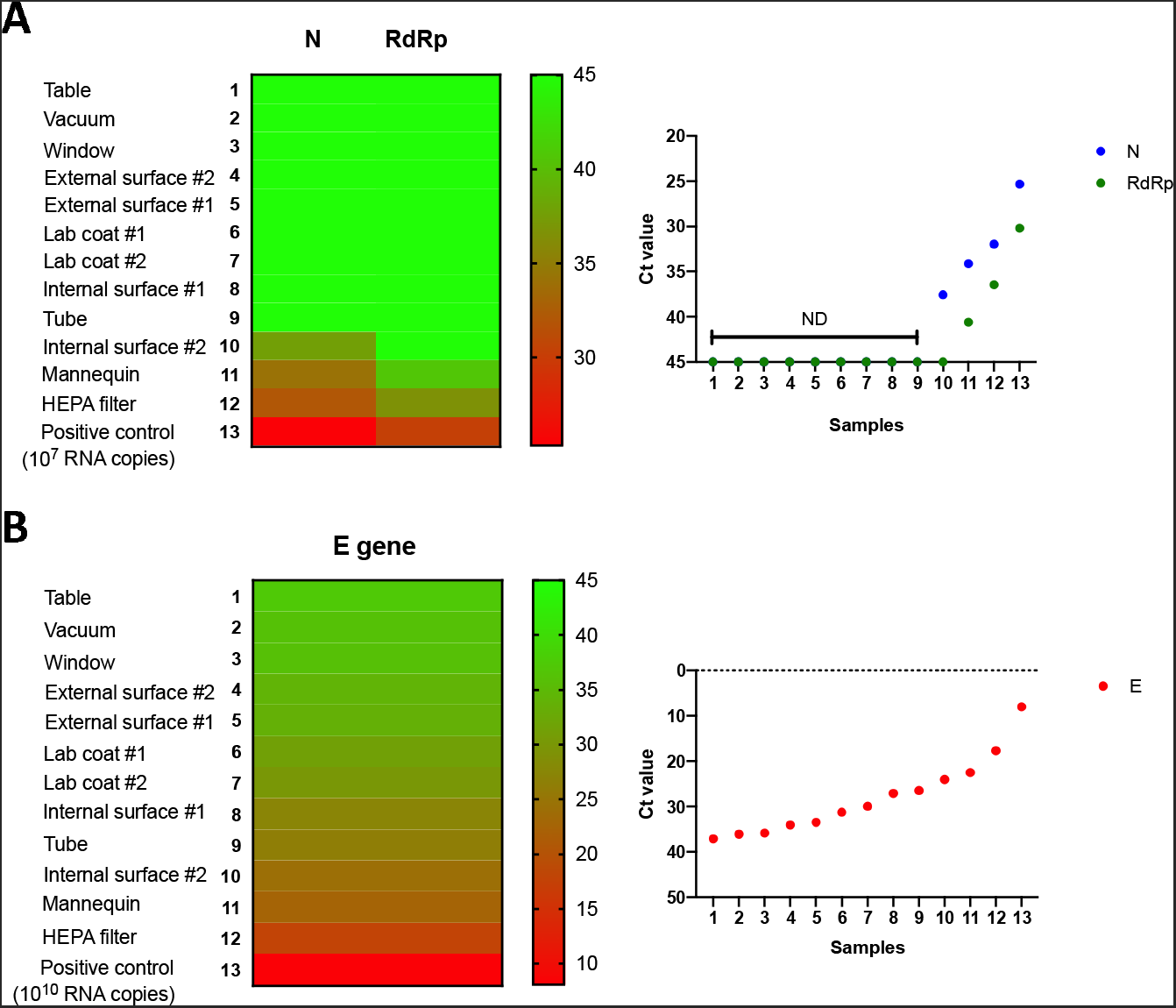
Evaluation of environment contamination by RT-PCR amplification of synthetic SARSCoV-2 RNA. (A) Detection of 10^7^ copies/ml of synthetic SARS-CoV-2 RNA by RT-PCR, of genes N and RdRP. (B) Detection of 10^10^ copies/ml of synthetic SARS-CoV-2 RNA by RT-qPCR, of E gene. Ct* values for each sample tested are represented as heatmaps (left) or scatter graph (right). ND = non detected after 45 amplification cycles. Locations of swab samples for RT-qPCR analysis. HEPPA filter: air influx position of the micro-particle filter used in the air suction, inside PIS/COVID-19; mannequin: inside PIS/COVID-19; internal surface # and #2: inner surface of the plastic cover, inside PIS/COVID-19; tube: exit position of the internal tube of PIS/COVID-19’s air suction system; lab coat #1 and #2: external surface of health professional’s disposable apron, after removing PIS/COVID-19; external surface #1 and #2: outer surface of the plastic cover, outside PIS/COVID-19; window: internal surface of hospital’s room window, next to the simulations; Vacuum: inner part of the hospital vacuum flask, connected to PIS/COVID-19’s air suction system; table: table next to the hospital’s bed where simulations occurred. *Ct = Cycle threshold, i.e. the amplification cycle in which fluorescence levels exceeds background threshold, being inversely proportional to the amount of nucleic acid present in the sample.

It is important to mention that caffeine has a molecular size in the order of Angstroms (10^−10^m) and a virus size in the order of nanometer (10^−9^m). However, the dissemination of SARS-CoV-2 occurs through aerosolization in particles that are in submicron (0.25 to 1.0 μm) or super micron (> 2.5 μm) sizes.^2^ Therefore, if PIS/COVID-19 is able to retain molecules smaller than viral particles, it should also be effective in preventing viral spread in the hospital environment. This presumption was confirmed when experiments were undertaken using synthetic SARS-CoV-2 RNA and monitoring results obtained by RTPCR analyses.

It is important to note that the simulations in this study occurred in a much more critical environment than real medical situations. During 900 minutes of the simulations, aerosolization of the caffeine solution occurred under an average pressure of 200 psi, equivalent to approximately 14,000 cm H_2_O. Pressure that is much higher than the one issued in a non-invasive ventilation that is of 30 cm H_2_O. In all simulations the internal environment of PISP/COVID-19 was saturated with the aerosolized solution, with a clear visual perception of droplet formation inside the cover.

In this experimental study, concentrations of synthetic RNA particles aerosolized inside PISP/COVID-19 were also chosen to simulate infecting conditions much higher than the virus infection concentrations described in clinical settings.^15–17^ Synthetic RNA virus in the concentration of 10^7^ copies/mL caused an aerosolization of 150 million virus particles, as 15 mL of the solution was aerosolized inside PISP/COVID-19. It simulated a condition with extremely high virus air concentration, not identified in publications describing virus air concentrations in clinical settings.^16, 17^ The concentration of 10^10^ synthetic RNA copies/mL aerosolized 150 billion viral particles, as 15 mL of the solution was used, simulating an inconceivable clinical condition.^16, 17^

Sars-CoV-2 concentrations in throat and sputum samples of infected patients ranged from 641 copies per mL to 1.34×10^11^ copies per mL, with a median of 7.99×10□/mL in throat samples and 7.52×10□/mL in sputum samples.^15^ The environment concentration of SARS-CoV-2 has less virus particles than identified in infected patient’s throat and saliva.^15–17^ Studies indicated that air virus concentrations, near hospitalized COVID-19 patients, ranged from 40×10^3^virus particles per air milliliter to maximum of 48.2×10^3^/mL of air next to infected patients receiving oxygen through a nasal cannula.^16, 17^ Mean room air virus concentrations in the study with highest values was of 28.6×10^2^/mL.^17^

Sars-CoV-1 infected patients have saliva concentrations of 7.08×10^3^ to 6.38×10^8^ copies per mL (median 9.92×10^4^ copies/mL).^18^Sars-CoV-1 nasopharyngeal concentrations ranged from 1.7×10^3^/mL to 3.4×10^7^/mL.^18^ Air concentration of Sars-CoV-1, near infected patients, ranged in concentrations of 1.1×10^1^ to 1.3×10^5^copies/mL.^19^The number of Sars-CoV-1 particles expelled per cough in infected patients is less than nasopharyngeal concentration.^18–20^ Sars-CoV-1 cough concentrations ranged from 900 particles to 30.2×10^4^ particles/mL/cough.^20^

Published data on air virus concentration near infected patients with Sars-CoV-1 and Sars-CoV-2 indicate total viral concentration that does not exceed the order of magnitude of 10^5^ particles per milliliter of air.^16,17, 19, 20^ This data confirms that the synthetic virus concentrations used in the present study (10^7^ copies/mL and 10^10^ copies/mL) are beyond the expected air concentrations near COVID-19 infected patients.

As air concentrations of Sars-CoV-1 and Sars-CoV-2 are similar.^16, 17, 19, 20^ Considering the 10^7^ copies/mL concentration of synthetic RNA particles used in this experimental simulations (150 million viral particles) and the maximum concentration of Sars-CoV-1 virus concentration per cough (30.2×10^4^particles/mL/cough)^20^, to reach the virus concentration used in the present study simulations, a patient would need to cough at least 496 times. In all the simulations, a total of 160 external sensor discs analyses were performed and there was no presence of caffeine found in all external sensors. Synthetic SARSCoV-2 RNA particles were used as an additional method to evaluate PISP/COVID-19’s ability to retain virus and protect the surrounding environment. Efficacy of PISP/COVID-19 was confirmed by detection and quantification of synthetic RNA by RT-PCR.

With these results, the PISP/COVID-19 is considered to be efficient in micro-particle retention in the simulated conditions. PISP/COVID-19 could be useful in indicating NIV and high-flow nasal cannula oxygen for hypoxemic COVID-19 patients without the need of having them in isolated, under laminar airflow and negative pressure rooms. It can also be of use to promote environmental protection by isolating infected patients under mechanical ventilation for up to 6 hours. It might protect health care professionals by lowering hospital air virus contamination. The aspiration of the internal environment of the PISP/COVID-19 in all simulations was continuous and it can be done through the hospital’s vacuum system or through a continuous suction pump.

Results of this experimental study proved PISP/COVID-19’s efficacy in situations related in this publication. Considering Sars-CoV-2 pandemic and urgent necessities of the Brazilian health system, to have PISP/COVID-19 available for clinical use, it had its approval by health authorities under the brand of BhioCOVID^®^.

## 4. Conclusions

The present experiment indicates that PISP/COVID-19 is effective in microparticle restraint when used in the conditions of NIV, high-flow nasal cannula oxygen and patient isolation under mechanical ventilation for periods up to 6 hours. There was no presence of the molecular marker outside the system. Synthetic SARS-CoV-2 RNA gradient inside and outside PISP/COVID-19 confirms its efficacy in viral retainment. In the current Sars-CoV-2 pandemic, it should be considered as a low-cost option that could diminish contamination of health professionals within the hospital environment by providing efficient COVID-19 patients isolation. It should be considered to guarantee environmental safety to provide NIV and high-flow nasal cannula oxygen in eligible COVID-19 patients, reducing the need of orotracheal intubation and mechanical ventilation.

## Data Availability

All data regarding molecular marker analysis with nuclear magnetic resonance spectroscopy and biological (synthetic SARS-CoV-2 RNA) evaluation using Reverse Transcription Polymerase Chain Reaction (RT-PCR) are available at any time.

